# Automated Deep Learning Pipeline for Callosal Angle Quantification

**DOI:** 10.1101/2025.08.18.25333901

**Authors:** Siavash Shirzadeh Barough, Murat Bilgel, Catalina Ventura, Lucas An, Ameya Moghekar, Marilyn S. Albert, Michael I. Miller, Abhay Moghekar

## Abstract

**BACKGROUND AND PURPOSE:** Normal pressure hydrocephalus (NPH) is a potentially treatable neurodegenerative disorder that remains underdiagnosed due to its clinical overlap with other conditions and the labor-intensive nature of manual imaging analyses. Imaging biomarkers, such as the callosal angle (CA), Evans Index (EI), and Disproportionately Enlarged Subarachnoid Space Hydrocephalus (DESH), play a crucial role in NPH diagnosis but are often limited by subjective interpretations. To address these challenges, we developed a fully automated and robust deep learning framework for measuring the CA directly from raw T1 MPRAGE and non-MPRAGE MRI scans.

**MATERIALS AND METHODS:** Our method integrates two complementary modules. First, a BrainSignsNET model is employed to accurately detect key anatomical landmarks, notably the anterior commissure (AC) and posterior commissure (PC). Preprocessed 3D MRI scans, reoriented to the Right Anterior Superior (RAS) system and resized to standardized cubes while preserving aspect ratios, serve as input for landmark localization. After detecting these landmarks, a coronal slice, perpendicular to the AC-PC line at the PC level, is extracted for subsequent analysis. Second, a UNet-based segmentation network, featuring a pretrained EfficientNetB0 encoder, generates multiclass masks of the lateral ventricles from the coronal slices which then used for calculation of the Callosal Angle.

**RESULTS:** Training and internal validation were performed using datasets from the Baltimore Longitudinal Study of Aging (BLSA) and BIOCARD, while external validation utilized 216 clinical MRI scans from Johns Hopkins Bayview Hospital. Our framework achieved high concordance with manual measurements, demonstrating a strong correlation (r = 0.98, p < 0.001) and a mean absolute error (MAE) of 2.95 (SD 1.58) degrees. Moreover, error analysis confirmed that CA measurement performance was independent of patient age, gender, and EI, underscoring the broad applicability of this method.

**CONCLUSIONS:** These results indicate that our fully automated CA measurement framework is a reliable and reproducible alternative to manual methods, outperforms reported interobserver variability in assessing the callosal angle, and offers significant potential to enhance early detection and diagnosis of NPH in both research and clinical settings.

## Introduction

Normal pressure hydrocephalus (NPH) is a potentially treatable neurological condition characterized by the triad of cognitive decline, gait disturbances, and urinary incontinence, as initially described by Hakim and Adams.^1^ Epidemiological studies reveal a wide range of prevalence estimates for NPH, but there is a consensus that it remains significantly underdiagnosed.^2,3^ This underdiagnosis is particularly concerning given the treatable nature of the disease.^4^ One of the major challenges in detecting NPH is its clinical overlap with other neurodegenerative diseases, such as Alzheimer’s Disease (AD), leading to frequent misdiagnoses.^5^ Furthermore, NPH is not often considered a primary differential diagnosis in elderly patients, despite its prevalence.^6^ This diagnostic gap highlights the need for enhanced tools and methodologies to increase awareness and improve the identification of NPH cases.^7^

Imaging biomarkers play a crucial role in the diagnosis of NPH.^8–10^ Measurements such as the Evans Index (EI)^11^, Callosal Angle (CA)^12^, and Disproportionally Enlarged Subarachnoid Spaces (DESH)^13^, provide objective evidence that supports clinical evaluation. Despite their diagnostic value, these imaging methods are often underutilized in clinical practice, primarily due to the perceived rarity of the condition and the time-intensive nature of manual image analysis. As a result, many potential cases may go undetected, emphasizing the necessity for automated, reliable, and efficient imaging tools to assist in the diagnosis of NPH.^14^ Implementing such tools can facilitate the identification of new patients and enable the retrospective analysis of previous scans, ensuring that individuals receive appropriate treatment.^15^

Recent advancements in artificial intelligence (AI)-based algorithms, coupled with the growing availability of medical imaging datasets and improved computational power, have paved the way for automating complex medical imaging tasks.^15^ In this study, we aim to develop a robust AI-based system to automatically measure the callosal angle from raw 1.5T and 3T MPRAGE MRI scans.^15^ By automating this critical measurement, our method seeks to improve the sensitivity and specificity of NPH diagnosis, reduce diagnostic delays, and assist clinicians in identifying potential cases, ultimately enhancing the care provided to patients with this treatable condition.

## Methods

### Data Collection

T1-weighted MRI data for training and internal validation were drawn from BLSA^16^ and BIOCARD. BLSA scans comprised: at 1.5 T, three GE Signa SPGR acquisitions (0.94 × 0.94 × 1.5 mm voxels; flip angle 45°; TE 5 ms; TR 35 ms; TI 0 ms) plus one Philips Intera 1.5 T SPGR system with identical parameters, and at 3 T, three Philips Achieva sagittal MPRAGE acquisitions (1 × 1 × 1.2 mm voxels; flip angle 8°; TE 3.2 ms; TR 6.5–6.8 ms; TI 0 ms). BIOCARD scans included a single 1.5 T GE Genesis Signa SPGR (axial; 1 × 1 × 2 mm voxels; flip angle 20°; TE 2 ms; TR 24 ms; TI 0 ms) and a single 3 T Philips Achieva MPRAGE (1 × 1 × 1.2 mm; flip angle 8°; TE 3.1 ms; TR 6.75 ms; TI 0 ms). For external validation, 216 T1-weighted scans were acquired at the Johns Hopkins Hydrocephalus Clinic on a Siemens Verio 3 T (0.78 × 0.78 × 0.80 mm voxels; flip angle 9°; TE 3.13 ms; TR 1800 ms; TI 900 ms). Additionally, to validate performance on non-MPRAGE data, 40 T1-weighted scans from the Johns Hopkins Hydrocephalus Clinic were included: 1.5 T spin-echo (SE) acquisitions (TR 736 ms; TE 8.7 ms; flip angle 90°; sequence variants SP and OSP) as well as 3 T T1 FLAIR Sag acquisitions (TR 2200 ms; TE 9.3 ms; flip angle 160°; scanning sequences SE and IR; sequence variants SK, SP, and MP). All images were anonymized by stripping private metadata and converted to NIfTI format using the dcm2niix Python package.

### Data Labelling

The 3D MRIs were anonymized and then annotated for the localization of the anterior commissure (AC) and posterior commissure (PC) coordinates using RilContour software.^17^ To accurately measure the callosal angle (CA), the coronal slice represented by the plane perpendicular to the line connecting AC to PC at the level of PC was extracted and saved as PNG images. These coronal slices were then segmented and annotated using Label Studio software to outline the right and left lateral ventricles. The callosal angle was measured according to the standard guidelines for CA calculation. All manual annotations and measurements were thoroughly verified by an experienced neurologist, thereby ensuring the highest standards of data accuracy and reliability.

### Landmark Detection Model

To automate the detection of PC and AC points that are crucial for the accurate measurement of the CA we used the BrainSignsNET^18^ model architecture which was pretrained on a large amount of data. We modified the structure of this model too be compatible with the detection of only AC and PC preserving the pretrained weights. The model was trained and finetuned on 80% of the manually annotated MRI scans from BLSA and BIOCARD using similar training process used for the training of the BrainSignsNET model described by its paper. Then the final trained model selected based on the performance of it on the 20% of the unseen data from BIOCARD and BLSA and was externally validated on the 216 scans from Johns Hopkins Hydrocephalus clinic.

### Ventricle Segmentation Model

For the segmentation task, we employed a UNet architecture^19^ integrating various pretrained backbone architectures. For the final model, the encoder was based on a pretrained EfficientNetB0, while the decoder was initialized with random weights. Preprocessed coronal MRI slices—resized to 224×224 pixels and standardized—served as inputs, and multiclass masks of lateral ventricles were the output of the model. The final convolution layer produces three‐channel logits per pixel (background, left ventricle, right ventricle) that are passed through a softmax to obtain class probabilities and then converted into the discrete multiclass segmentation mask.

A dual loss function was used to optimize performance: combining the Dice Score Coefficient (DSC) with the Hausdorff Distance (HD). The HD component was specifically incorporated due to its enhanced sensitivity to the segmentation mask borders, which are of critical importance in this project.

To further improve model generalizability, data augmentation techniques—including rotation, flipping, random blurring, and the addition of noise—were applied. The model was initially trained on 80% of the images extracted from the BLSA and BIOCARD cohorts, with the remaining 20% reserved for internal validation. The data split was performed based on patient IDs to prevent data leakage. Finally, the best performing model was externally validated using data from the Johns Hopkins Hydrocephalus Clinic to assess its performance on unseen clinical datasets.

### Callosal Angle Calculation

To compute the callosal angle (CA) the following steps were applied, as illustrated in Figure 1:

**Figure 1.**
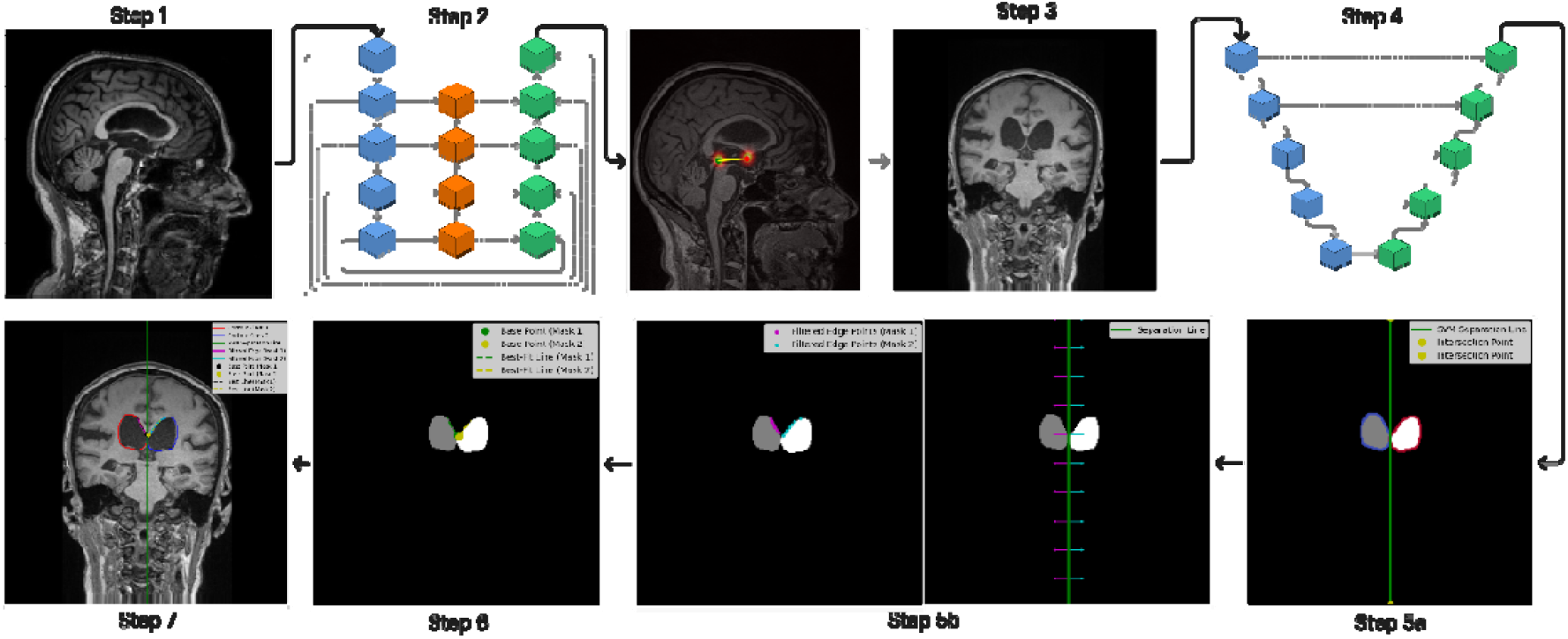
Overview of the automated processing pipeline for callosal angle measurement from raw MRI data: 3D MRI scans are first reoriented to the RAS coordinate system, resized to 128×128×128 cubes (preserving aspect ratio), and transformed using standard scaling (Step1). The anterior and posterior commissures (AC and PC) are then automatically detected using the trained BrainSignsNET model (Step 2), and a coronal slice—perpendicular to the AC–PC line at the level of the PC—is extracted (Step 3). Segmentation masks of the lateral ventricles are generated using UNet model (Step 4), and their marginal contours are delineated. Subsequently, an SVM algorithm computes a separating line with maximum margin between the two ventricles (Step 5a), after which a search algorithm projects perpendicular rays from this line to identify the initial collision points that represent the medial walls (Step 5b). These contact points, satisfying a predefined threshold, serve as base points from which a line representing the medial wall is derived (Step 6). Finally, vectors from these medial wall lines are used to calculate the callosal angle Step 7.

1. **MRI Preprocessing:** 3D MRI scans were reoriented to Right Anterior Superior (RAS), resized to 128,128,128 cubes preserving the aspect ratio of the original scans and transformed using standard scaler.
2. **Landmark Detection:** The AC and PC will be detected from pre-processed 3D MRI scans using the trained BrainSignsNET model.
3. **Coronal Slice Extraction:** The coronal slice represented by the plane perpendicular to the line connecting AC to PC at the level of PC will be extracted.
4. **Segmentation and Contouring:** Segmentation masks of the lateral ventricles will be extracted, and marginal contours will be identified using a contouring algorithm.
5. **Identification of the medial walls of the lateral ventricles: a:** Using Support vector Machine (SVM) algorithm a line separating 2 ventricles with maximum margin will be calculated then **b**: using a specific search algorithm which sends perpendicular lines from the separating line (rays) and calculate first collision point on each ventricle which represent the medial wall of lateral ventricles.
6. **Identification of linear representation of medial walls of the lateral ventricles:** Points of contact (closest point of each ventricle to the other) meeting a defined threshold between the two ventricles were identified and marked as the base points for each ventricle. Then, using a search method a line starting from each base point that crosses most points of the medial wall of each side will be identified.
7. **Angle Calculation:** Vectors derived from the lines representing medial wall of lateral ventricles were used to calculate the angle between them, representing the callosal angle.

### Statistical Analysis and Evaluation

Numerical demographic data were summarized as mean and standard deviation, while categorical demographic data were reported as counts and percentages.

The performance of the landmark detection model was quantified by calculating the three-dimensional Euclidean Distance between the predicted and manually annotated landmarks. In addition, we computed the Percentage of Correct Key Points (PCK@10), whereby predictions with an error less than 10% of the Euclidean distance between the anterior and posterior commissures (AC-PC) were classified as correct. Predictions that exceeded this error threshold were deemed incorrect, and this metric was subsequently used to report the model’s overall accuracy in predicting anatomical landmarks.To assess the pitch correction—defined as adjusting the 3D MRI scan such that the AC-PC line is aligned parallel to the horizontal plane in both sagittal and axial axes (ensuring the coronal plane is perpendicular to the AC-PC line)—we calculated the angular difference between two lines: one formed by the manually annotated AC and PC points, and the other determined by our predictive method. This angular difference served as the error metric for the pitch correction. The method’s performance was quantified by computing the Mean Absolute Error (MAE). In addition, a Bland-Altman analysis was conducted to visually assess the agreement between manual annotations and predicted alignments. The results of this analysis are reported by both the bias and the 95% limits of agreement.

We evaluated the performance of the segmentation model using the Mean Dice Similarity Coefficient (MDSC) and the Mean Hausdorff Distance (MHD). These metrics quantified the overlap and boundary agreement between the predicted (*A*_*i*_) and ground truth (*B*_*i*_) segmentations, respectively. MDSC and MHD were calculated using the following formulas:

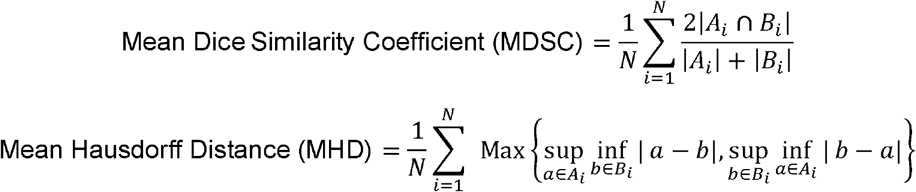

The final callosal angle measurements will be assessed using MAE and Pearson correlation analysis to compare predicted and ground truth values. A threshold of 90° will be employed to classify patients into normal or abnormal categories based on both predicted and annotated callosal angle measurements.

External validation will be conducted on a clinically acquired dataset from the Hydrocephalus Clinic at Johns Hopkins Bayview Medical Campus. This dataset comprises patients predominantly diagnosed with hydrocephalus, with a significant proportion exhibiting positive callosal angle measurements. This ensures the robustness and generalizability of the proposed method.

To assess the effect of localization and orientation errors on callosal‐angle (CA) measurements, we introduced controlled perturbations of the PC landmark and plane orientation. Specifically, we shifted the PC position in 1 mm increments up to ±10 mm along the anterior–posterior axis and rotated the plane normal in 1° increments up to ±10° about both the sagittal and axial axes. After each perturbation, the plane was re‐extracted, segmented, and the CA re‐measured. By directly comparing each perturbed measurement to the original, unperturbed CA, we quantified how incremental spatial or angular deviations propagate into CA measurement bias.

Statistical significance was defined as two-tailed p < 0.05.

### Training and Validation Setup

All processing and model training were conducted on a high-performance workstation equipped with dual Nvidia RTX 4090 GPUs, 128 GB RAM, and an AMD Threadripper Pro WRX80 CPU. Training was performed for 100 epochs or until convergence, using the AdamW optimizer with a weight decay of 0.02.

## Results

### Demographics

MRI scans analyzed in this study originated from three cohorts summarized in Table 1: the Baltimore Longitudinal Study of Aging (BLSA), BIOCARD, and Johns Hopkins Clinic. Overall, the study included a total of 1,127 individuals with 2,123 MRI scans. The BLSA cohort (n=516) had a mean age of 69.31 years (SD=15.55). The BIOCARD cohort (n=395) contributed the largest number of scans (1,391 scans) and had an average age of 64.22 years (SD=11.02), also utilized for training and internal validation. The Johns Hopkins Clinic cohort (n=216), with a mean age of 71.99 years (SD=8.09), provided external validation scans with a Mean Evans Index measurement of 0.37 (SD=0.05).

**Table 1.**
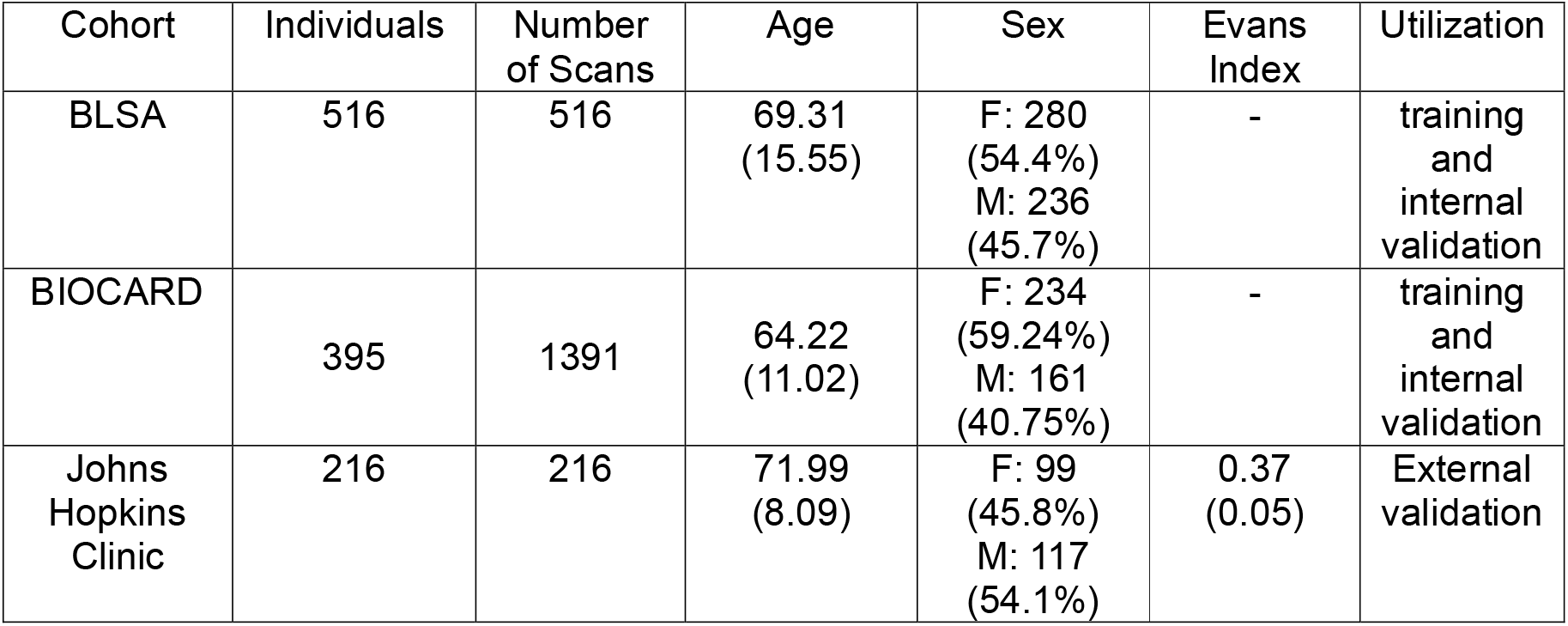
summarizes the demographic characteristics of participants included in the training and validation datasets. In this context, “Age” refers to the participants’ age at the time of MRI acquisition. The table also details gender distribution, the number of scans per dataset, the total number of participants, and the mean Evans Index with its standard deviation (mean ± SD).

### Landmark detection and MRI pitch correction

The final evaluation of the trained BrainSignsNET model for detecting the 3D coordinates of anatomical landmarks demonstrated high accuracy and robustness. Specifically, internal validation yielded a median error (MED) of 1.8 mm for the AC and 1.57 mm for the PC, External validation showed similar performance, with MEDs of 1.81 mm for AC and 1.92 mm for PC. Moreover, each predicted landmark was within 10 mm of the corresponding annotated landmark, achieving a PCK@10 value of 100% across all evaluations.

The performance error of the pitch correction is visualized in Figure 2a These results indicate an overall pitch correction characterized by 95% limits of agreement ranging from −4.4° to +3.92°, with a bias of −0.24°. Additionally, the Mean Absolute Error (MAE) was 1.57° with a standard deviation of 1.44°. The pitch angle using predicted landmarks had a significant correlation with the manually annotated pitch angle (r = 0.88, p-value < 0.001) Figure 2b.

**Figure 2.**
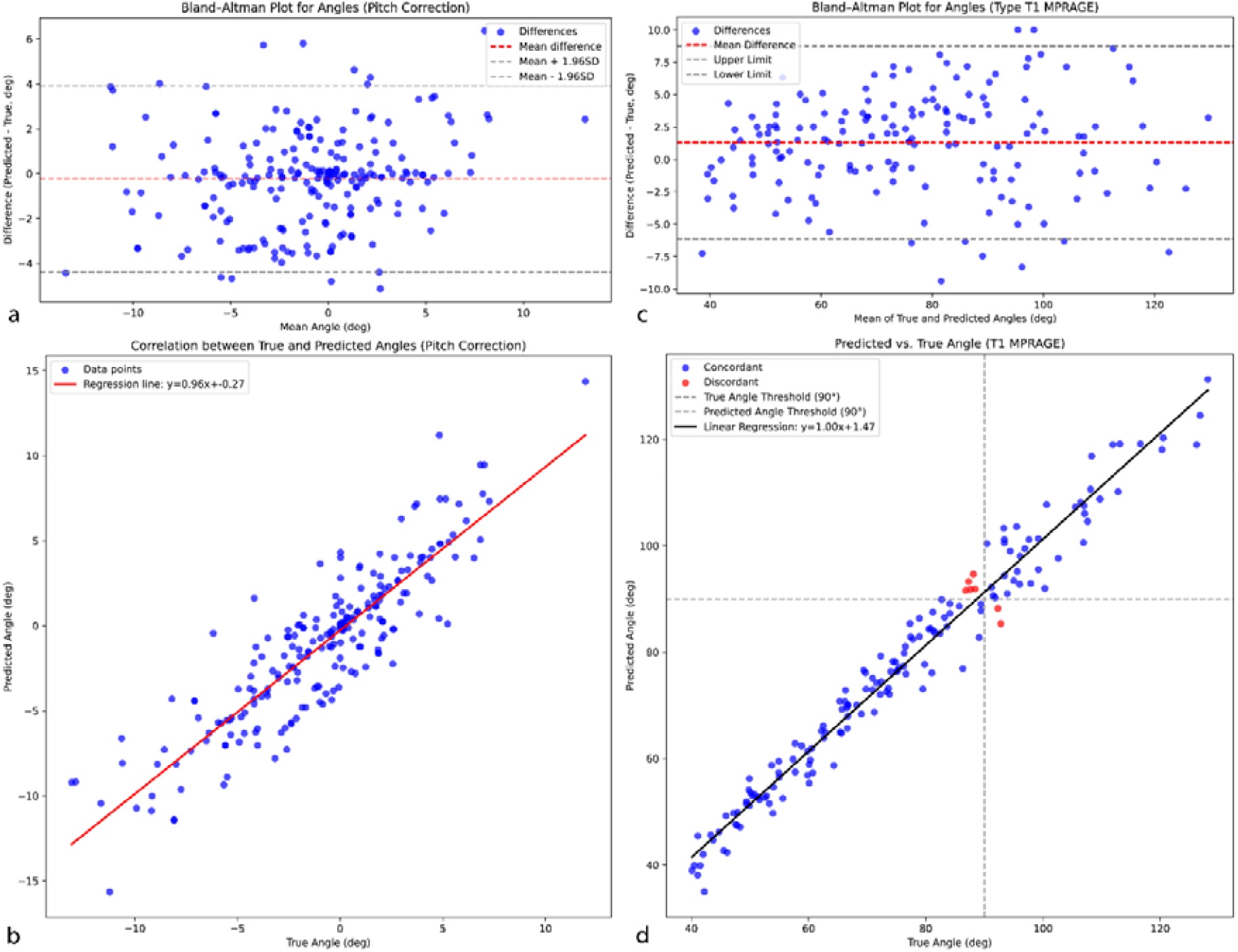
Overview of the performance evaluation of the automated framework. Panels (a)–(b) and (c)–(d) illustrate the performance of AC-PC angle (pitch) and callosal angle estimation, respectively. Panel (a) shows the Bland-Altman plot assessing the performance error of the pitch correction procedure relative to manual annotations, while panel (b) illustrates the correlation between the pitch angles obtained from the predicted landmarks and those from manual annotations. Panel (c) presents the Bland-Altman analysis comparing the overall automated callosal angle (CA) measurements to the manual CA measurements, and panel (d) displays the correlation between the predicted and manually measured CA values. Additionally, panel (d) indicates in red the CA measurements that would be incorrectly classified as narrow versus wide based on a 90° threshold.

### Segmentation Models

Among the evaluated segmentation architectures, the UNet model equipped with a pretrained EfficientNetB0 encoder demonstrated superior overall performance (compared with the performance of simple UNet architecture with overall Dice score of 79.21% and UNet with DenseNet121 as backbone with overall Dice score of 89.18%). On the internal validation dataset, the model achieved an overall Dice score of 92.49% and a mean Hausdorff distance (MHD) of 1.29 mm. Detailed analysis revealed that the right and left ventricles achieved Dice scores of 92.31% and 92.68% with MHDs of 1.38 mm and 1.21 mm, respectively. In the external validation dataset, the model maintained its strong performance with an overall Dice score of 94.31% and an MHD of 1.64 mm; specifically, the right and left ventricles attained Dice scores of 93.13% and 95.49%, respectively, with corresponding MHDs of 1.68 mm and 1.60 mm.

### Callosal Angle Measurement Performance

As demonstrated in Figure 2c, overall performance evaluation of the proposed algorithm in measurement of CA from 3D raw MRI showed a MAE of 2.95 (SD=1.58) degrees between the predicted CA and the manually measured CA. The Bland-Altman analysis demonstrated that the overall automated CA measurement yielded 95% limits of agreement ranging from −6.17° to +8.75°, with a bias of 1.31°. The analysis showed a significant correlation between predicted and manually annotated CA with a correlation coefficient of 0.98 and P-value < 0.001 (Figure 2d). Moreover, considering a threshold of 90° for classifying narrow versus wide callosal angle, out of 216 scans in the external validation dataset, only 7 scans were classified incorrectly.

To further evaluate the clinical applicability of our methodology for assessing the Callosal Angle (CA), as illustrated in Supplementary Figure 2, we applied the method to 40 non-MPRAGE scans. The predicted CA values were compared to the manual measurements obtained from the corresponding MPRAGE scans. This comparison yielded a mean absolute error (MAE) of 3.3° ± 1.61°. A subsequent Bland-Altman analysis indicated that the 95% limits of agreement ranged from −9.93° to +10.56°, with a bias of 0.31°. (Supplementary Figure 2a) Moreover, correlation analysis demonstrated a highly significant relationship between the predicted and manually annotated CA values, with a correlation coefficient of 0.98 (p < 0.001; Supplementary Figure 2b).

### Error and Bias analysis

Figure 3 illustrates that neither the pitch correction error nor the callosal angle prediction error exhibited statistically significant correlations with age or the Evans index. Additionally, no significant differences were found between genders for either error metric (all p-values > 0.05).

**Figure 3.**
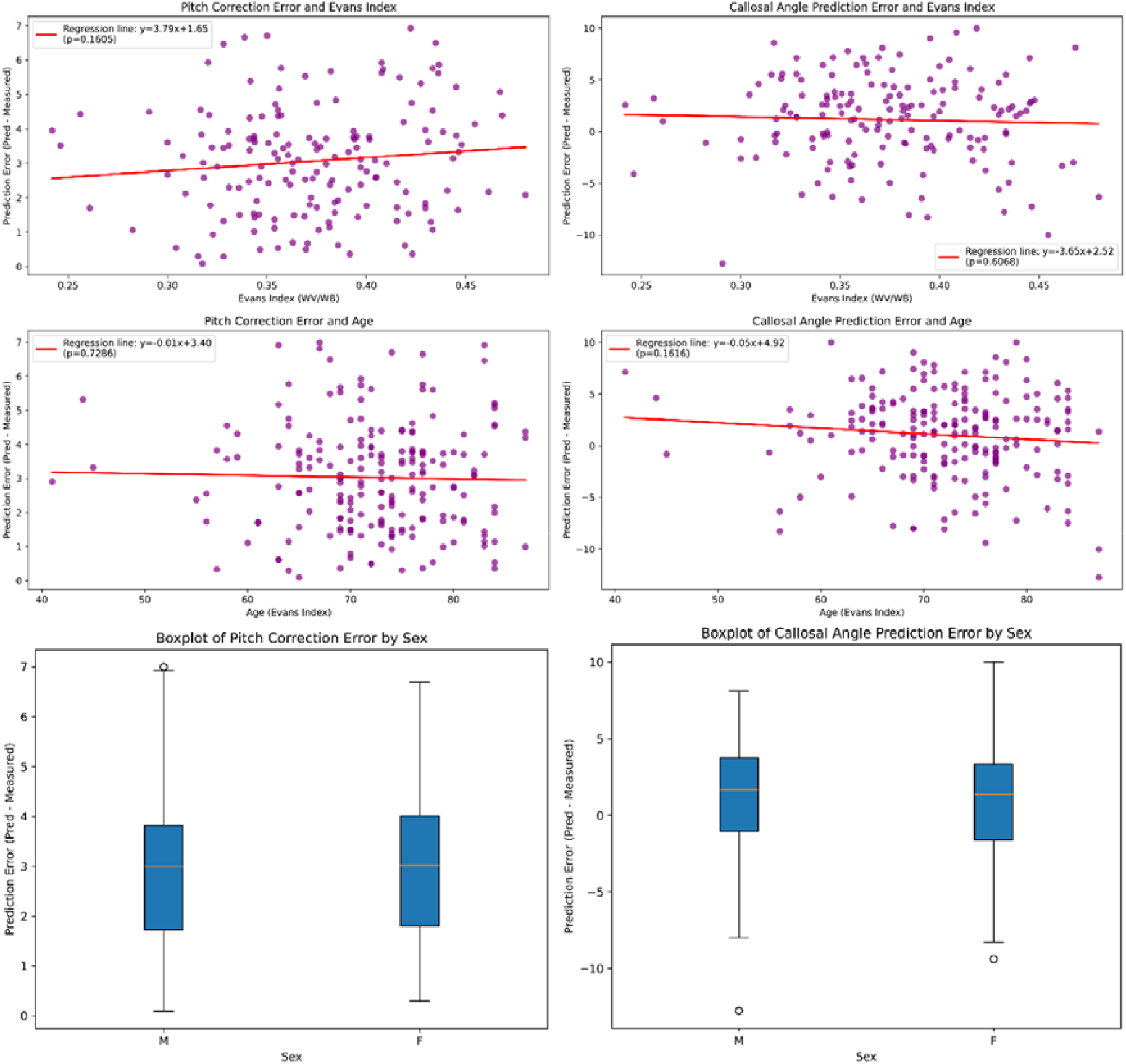
Evaluation of the error differences in the automated framework stratified by clinical and demographic parameters. Panels (a) and (b) are scatter plots showing the correlation between pitch correction errors and the Evans Index and age, respectively, while panel (c) is a box plot depicting the differences in pitch correction errors by gender. Similarly, panels (d) and (e) are scatter plots illustrating the relationship between callosal angle measurement errors and the Evans Index and age, respectively, with panel (f) presenting a box plot of callosal angle measurement errors stratified by gender.

In the sagittal‐offset analysis (Figure 4a), introducing a 5° pitch error increased the CA measurement error to approximately 6.13 ± 4.45° (clockwise) and 4.77 ± 3.62° (counterclockwise). When the pitch error was doubled to 10°, the measurement error rose further to 9.21 ± 6.64° (clockwise) and 7.63 ± 6.20° (counterclockwise). In the axial‐offset analysis (Figure 4b), a 5° rotation increased the callosal‐angle error to 3.26 ± 2.67° (clockwise) and 3.91 ± 3.10° (counterclockwise). A 10° axial error showed 4.76 ± 4.20° (clockwise) and 4.74 ± 3.94° (counterclockwise). Finally, shifting the PC landmark posteriorly by 5 mm increased the CA error to 10.60 ± 6.99°, and a 5 mm anterior shift yielded 12.95 ± 8.26° (Figure 4c). At the extremes (±10 mm), posterior displacement produced 16.58 ± 8.91° of error, while a 10 mm anterior shift produced 25.63 ± 14.45°. In all three perturbation experiments, each nonzero deviation was significantly different from baseline (paired t‐tests, p ≪ 0.001). (Supplementary Figure 1)

**Figure 4.**
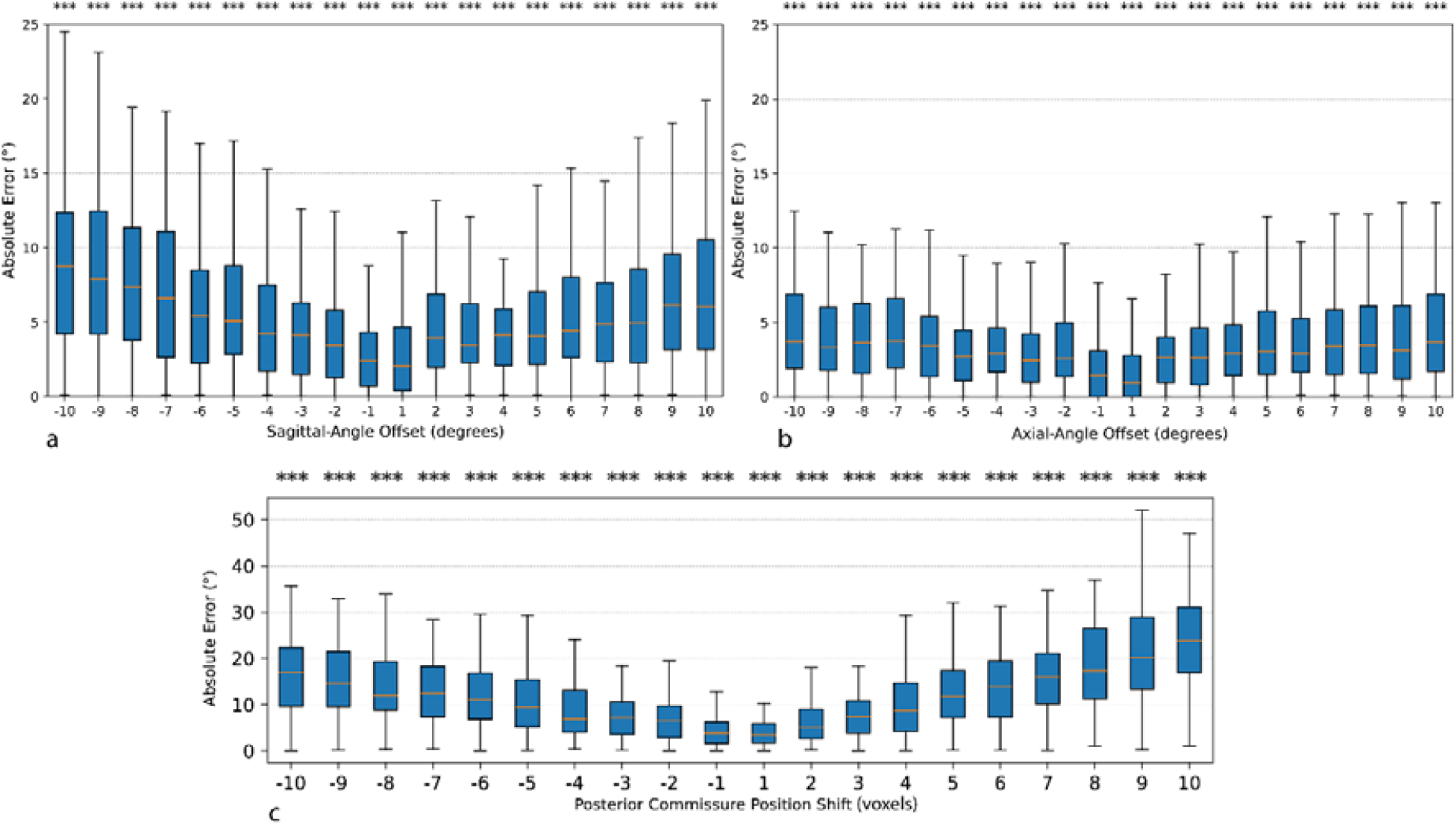
Boxplots of absolute callosal‐angle (CA) measurement error as a function of perturbation magnitude for three sources of potential bias: sagittal‐plane angle offsets (a), axial‐plane angle offsets (b), and posterior commissure (PC) landmark shifts (c). For each subplot, the x‐axis indicates the magnitude of the perturbation (in voxels (equals to 1mm) for PC shifts; in degrees for sagittal and axial rotations), and the y‐axis shows the resulting absolute CA error (°) compared to the unperturbed baseline. Asterisks above each box denote the level of statistical significance (^*^ p < 0.05; ^**^ p < 0.01; ^***^ p < 0.001) from paired t‐tests against the zero‐perturbation condition.

## Discussion

In this study we introduce a fully automated framework for the measurement of the callosal angle (CA) from raw MRI data according to the CA measurement method described by Ishii et al.^20^, offering an objective and reproducible approach to quantifying this clinically significant index. The framework synergistically integrates two deep learning models designed to perform complementary tasks. The first model detects key anatomical landmarks in three-dimensional MRI data—specifically, the anterior and posterior commissures—while the second model executes two-dimensional ventricular segmentation to delineate accurate ventricular contours. By combining these techniques, the framework establishes an robust methodology for CA measurement, significantly reducing reliance on manual, error-prone procedures. Extensive evaluations on both internal and external validation datasets—comprising research-grade as well as clinically acquired MRI scans— demonstrate the method’s high accuracy and generalizability. The framework not only exhibits robust performance on previously unseen clinical T1 MPRAGE scans for CA calculation but also maintains acceptable performance on non-MPRAGE clinical scans, which are commonly used in clinical settings for brain evaluation. Furthermore, bias analysis indicates that age, sex, and the severity of hydrocephalus (as quantified by the Evans Index) are not associated with model performance, thereby ensuring its applicability especially to patients with a wide range of ventricular volumes.

Previous studies have proposed methods to evaluate MRI metrics related to normal pressure hydrocephalus (NPH), including the Evans Index^21,22^, callosal angle^15^, and DESH^23^, as well as approaches for direct NPH diagnosis from MRI scans. Many of these methodologies rely on either manual procedures or automated segmentation tools, such as FreeSurfer^24^ for three-dimensional brain segmentation, which often lack generalizability when applied to clinically acquired MRI scans, particularly those deviating from standard protocols or involving atypical cases like severe ventricular enlargement or hydrocephalus. In these contexts, segmentation errors become more pronounced and critical anatomical landmarks, such as the anterior and posterior commissures, are not properly identified, leading to discrepancies in indices such as the CA. Furthermore, studies have demonstrated that even minor deviations in pitch correction—errors of less than 10° relative to the AC-PC plane—can significantly alter CA measurements.^25^ In our study, we similarly confirmed that small misalignments or landmark‐localization errors produce significant CA bias. For instance, modest sagittal‐plane rotation or axial tilt each introduced several degrees of angular error, and shifting the PC landmark (coronal slice location) by only a few millimeters resulted in substantial deviations in CA measurement. These findings underscore that precise, consistent localization and alignment are critical to obtaining valid callosal‐angle measurements. A similar concept applies to other MRI-derived measurements, such as the Evans Index. The EI is calculated on the axial plane that captures the maximal width of the anterior horns of the lateral ventricles—a plane that must be perpendicular to the AC-PC line. Consequently, accurate detection of these anatomical landmarks is imperative for the reliable computation of various MRI-driven indices.

In this study, we developed a robust and externally validated deep learning framework for callosal angle (CA) measurement that leverages a combination of landmark detection and segmentation algorithms to extract precise anatomical landmarks and delineate ventricular structures. This integrated approach adheres closely to standardized methodologies, ensuring a transparent CA measurement process that facilitates the visualization and identification of potential errors, thereby enhancing clinical utility. Our framework demonstrated accuracy comparable to manual measurements and broad applicability across both research and clinical datasets, including MRI scans from patients with markedly enlarged ventricles as our analysis showed no significant correlation between the error related to pitch correction and CA measurements and severity of hydrocephalus evaluated by EI. Moreover, all models performed well in both internal and external validation datasets, ensuring the robustness and generalizability of the method.

Furthermore, our method exhibits superior performance relative to previous automated CA measurement approaches. While a prior study reported 95% limits of agreement ranging from –29.99° to +21.98° and interobserver variability among radiologists with 95% limits of agreement from –14.95° to +7.39°, our framework achieved markedly lower variability, with 95% limits of agreement from –6.14° to +8.79°. These findings underscore the enhanced reliability and clinical relevance of our approach, positioning it as a reliable solution for both clinical diagnostics and research applications in NPH-related MRI assessments.^15^

Despite the promising results, this study has certain limitations. First, the datasets used for training and internal validation primarily included scans from research cohorts and a single clinical center, which may not fully capture the variability of imaging protocols and patient populations encountered in broader clinical practice. Future work should focus on expanding the training datasets to include more diverse populations and imaging settings. Second, while the model demonstrated excellent performance in detecting abnormal CA values, its integration into a comprehensive diagnostic framework for NPH requires further validation in prospective clinical studies. Third, this framework was only trained and validated on T1-weighted MRI scans. Future research should also explore the extension of this pipeline to include additional imaging biomarkers, such as the Evans Index and DESH patterns, to provide a more comprehensive diagnostic tool for NPH. Additionally, adding the capability to analyze other types of imaging including CT scans and other MRI sequences would make this model more useful in both clinical and research settings.

## Conclusion

This study presents a robust AI-based system for the automated measurement of the callosal angle from raw MRI scans. By achieving high accuracy and reliability across key diagnostic tasks, the proposed method has the potential to improve the sensitivity and specificity of NPH diagnosis, reduce diagnostic delays, and assist clinicians in identifying treatable cases. These findings mark a significant step toward the development of AI-driven tools for enhancing the care of patients with NPH and other neurological conditions.

## Data Sharing Statement

The complete codebase for Automated Callosal Angle Measurement, including all scripts and implementations of the BrainSignsNet, 2D Ventricular Segmentation, and CA Measurement code, is publicly available at https://github.com/SiavashShirzad/AutomatedCallosalAngle. The pretrained model weights used in this study will be provided upon reasonable request from the corresponding author and are planned to be released publicly in a future update.

## Acknowledgments

This research was supported in part by the Intramural Research Program of the National Institutes of Health (NIH). The contributions of the NIH author(s) were made as part of their official duties as NIH federal employees, are in compliance with agency policy requirements, and are considered Works of the United States Government. However, the findings and conclusions presented in this paper are those of the author(s) and do not necessarily reflect the views of the NIH or the U.S. Department of Health and Human Services.

