## Supplementary Figure 1 for "Automated Deep Learning Pipeline for Callosal Angle Quantification"

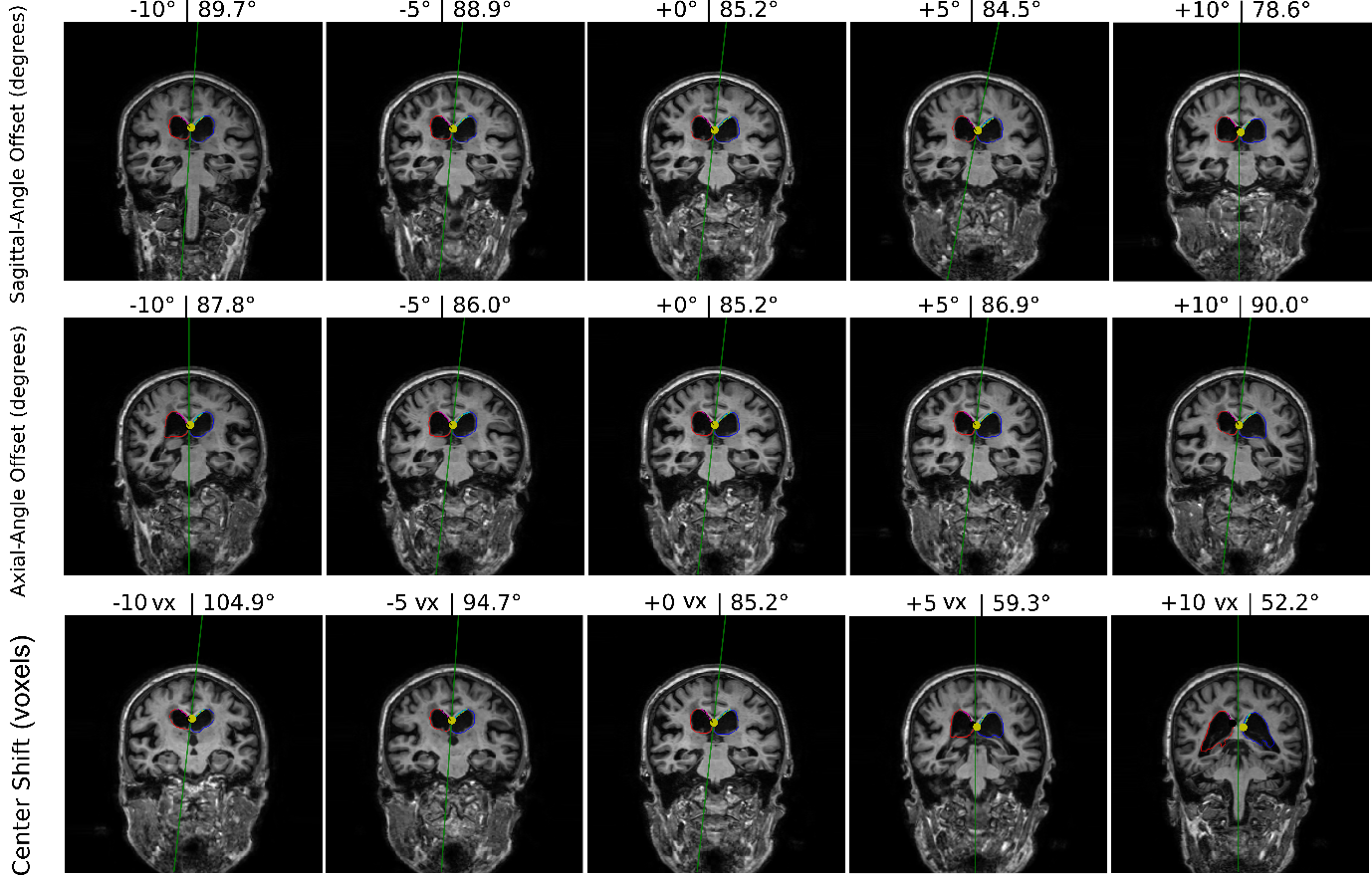

Supplementary Figure 1. Coronal‐plane for one subject under three perturbation types: sagittal‐plane rotations, axial‐plane rotations, and PC landmark shifts. Each row shows five steps (−10, −5, 0, +5, +10) of bias in angles (degree) and location (mm). As well as calculated Callosal Angle (degree).
