## Supplementary Figure 2 for "Automated Deep Learning Pipeline for Callosal Angle Quantification"

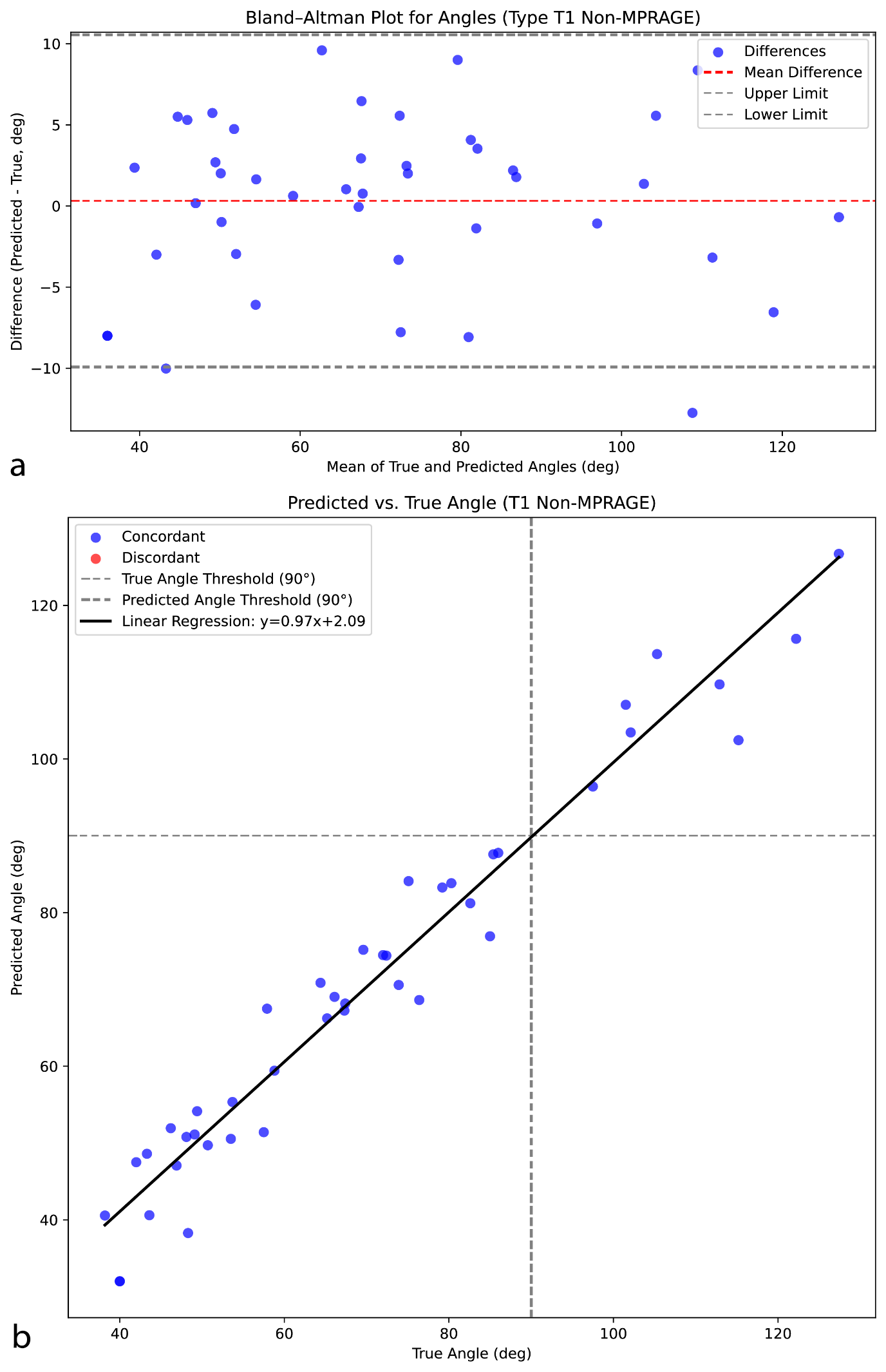

Supplementary Figure2. Overview of the evaluation of callosal angle (CA) measurements on non-MPRAGE scans. Panel (a) displays a Bland-Altman plot comparing the predicted CA values with the manual measurements obtained from corresponding MPRAGE scans. Panel (b) illustrates the correlation between the predicted and manually annotated CA values via a scatter plot.
